# Does sub-Saharan Africa truly defy the forecasts of the COVID-19 pandemic? Response from population data

**DOI:** 10.1101/2020.07.06.20147124

**Authors:** Dongmo Christophe Fokoua-Maxime, Monique Amor-Ndjabo, Amandus Ankobil, Momah Victor-Kiyung, Steve Franck-Metomb, Simeon Pierre Choukem

## Abstract

**Introduction:** Since its identification, the COVID-19 infection has caused substantial mortality and morbidity worldwide, but sub-Saharan Africa seems to defy the predictions. We aimed to verify this hypothesis using strong statistical methods.

**Methods:** We conducted a cross-sectional study comparing the projected and actual numbers as well as population proportions of COVID-19 cases in the 46 sub-Saharan African countries on May 1^st^, May 29^th^ (4 weeks later) and June 26^th^ (8 weeks later). The source of the projected number of cases was a publication by scientists from the Center for Mathematical Modeling of Infectious Diseases of the London School of Hygiene & Tropical Medicine, whereas the actual number of cases was obtained from the WHO situation reports. We calculated the percentage difference between the projected and actual numbers of cases per country. Further, “N-1” chi-square tests with Bonferroni correction were used to compare the projected and actual population proportion of COVID-19 cases, along with the 95% confidence interval of the difference between these population proportions. All statistical tests were 2-sided, with 0.05 used as threshold for statistical significance.

**Results:** On May 1^st^, May 29^th^ and June 26^th^, respectively 40 (86.95%), 45 (97.82%) and 41 (89.13%) of the sub-Saharan African countries reported a number of confirmed cases that was lower than the predicted number of 1000 cases for May 1^st^ and 10000 for both May 29^th^ and June 26^th^. At these dates, the population proportions of confirmed Covid-19 cases were significantly lower (p-value <0.05) than the projected proportions of cases. Across all these dates, South-Africa always exceeded the predicted number and population proportion of COVID-19 infections.

**Conclusion:** Sub-Saharan African countries did defy the dire predictions of the COVID-19 burden. Preventive measures should be further enforced to preserve this positive outcome.

## Introduction

On December 12^th^, 2019, an epidemic caused by the coronavirus disease 2019 (COVID-19) emerged in Wuhan, China [1]. The epidemic developed rapidly and spread to many countries. On March 11^th^, 2020, the World Health Organization (WHO) declared the COVID-19 outbreak a pandemic [2].

The first case of COVID-19 on the African continent was detected in Egypt on February 12^th^, 2020 [3,4]. Since then, the pandemic has substantially progressed and as of July 4^th^, 2020, African countries reported a total of 342 415 confirmed cases and 6 628 deaths [5]. The pandemic did catch the continent off guard and has since obliged political leaders to take important decisions constantly. The rapid progression of the sanitary crisis constrains those stakeholders to make these decisions based on information of the highest accuracy possible, with regards to the number of lives at stake. Since history is the best teacher, historic data are usually utilized as reference for decision making purposes. However, COVID-19 is a novel disease and thus no historic data exist on the pandemic. Therefore, to raise alarms and build public health strategies, public health officials can only rely on predictive models. These models are informed by the existing knowledge about population features and habits as well as the growing evidence on the COVID-19 infection.

Sub-Saharan African populations are very dense and one of the key characteristics of their social life is the presence of quasi-constant human contacts (handshakes, hugs, accolades), which make it very difficult to apply social distancing. Furthermore, potable water is not readily available, and poverty prevents most of these populations from affording either the face masks or the materials necessary to implement and enforce ideal hygiene sanitation measures. Finally, a great percentage of sub-Saharan African subjects rely on a daily income to sustain a living; as such, it is not possible to maintain confinement measures for long periods of time. Based on these facts, dire predictions were made about the evolution of the COVID-19 infection in Africa. A statistical model built by Pearson et al. (2020), from the Center for Mathematical Modeling of Infectious Diseases of the London School of Hygiene & Tropical Medicine (LSHTM) predicted that most African countries would report 1000 cases by May 1^st^ and 10,000 cases few weeks later [6].

The World Health Organization alerted on April 17^th^, 2020 that Africa would become the next epicenter of the COVID-19 pandemic [7,8]. As the pandemic has evolved, several press releases have highlighted that sub-Saharan African countries seemed to have defied the odds of such predictive models [9–11]. Although seemingly positive, such news have left most researchers dissatisfied because there has not been any scientific appraisal of the difference between these predictions and the actual data. In effect, to the best of our knowledge, there exists no quantitative assessment of the difference between the actual and projected number of COVID-19 cases in sub-Saharan Africa. Further, no studies have attempted to determine if there is a statistically significant difference between the projected and actual prevalence of the COVID-19 infection and explain any pattern. Our research therefore aimed to use the predictions made by Pearson et al. (2020), to (1) determine the percent difference between the actual and projected number of COVID-19 cases in sub-Saharan African countries at the indicated dates, (2) assess if there is a statistically significant difference between the projected and actual population proportion of COVID-19 at these dates and (3) discuss any pattern.

## Methods

### Data sources

We conducted a cross-sectional study analyzing the projected and actual number as well as population proportion of COVID-19 cases in the 46 countries of sub-Saharan Africa. The date of May 1^st^ was the date indicated by Pearson et al. (2020), [6] to describe the day at which, on average, most African countries were projected to have reached 1000 cases. They further stated that most African countries would have reached 10000 cases “couple of weeks” after May 1^st^. Because the timeline to reach the 10000 cases was not specifically indicated, we choose to make comparisons with the actual number of confirmed cases reported on May 29^th^ (4 weeks after May 1^st^) and June 26^th^ (8 weeks after May 1^st^).

Pearson et al. (2020), obtained these predictions from robust statistical models built around established epidemiologic parameters like the average number of additional cases that each case produces [12] and the average time between the onset of a case and the onset of a subsequent case infected by that case [13]. The starting date for these projections was the date when each country had 25 confirmed cases reported in the World Health Organization Situation Reports (SITREPs); for countries who had not reached this number by March 24^th^, the projections were made from March 25^th^.

To perform the comparisons, we extracted data on the actual number of confirmed COVID-19 cases from the SITREPs of May 1^st^ [14], May 29^th^ [15] and June 26^th^ [16]. The 2019 total population estimates were obtained from the United Nations World Population Prospects 2019 [17].

Our study was exempt from Institutional Review Board approval since we used data already collected.

### Data Analysis

We first determined the percentage difference between the projected and actual number of COVID-19 cases by country on May 1^st^, May 29^th^ and June 26^th^. To do so, we calculated the difference between the actual and projected number of cases, divided it by the projected number of cases and converted the results to percentages. Further, we computed the projected and actual population proportion of COVID-19 infection by country on May 1^st^, May 29^th^ and June 26^th^; we did it by dividing the total number of confirmed cases reported in the SITREPs by the total population count of each country indicated the United Nations World Population Prospects 2019. Thereafter, we compared the values of the projected and actual proportions of COVID-19 cases using the two-sided “N-1” Chi-squared test as recommended by Campbell [18] and Richardson [19]. Further, we performed Bonferroni correction to reduce the risk of type 1 error while performing multiple statistical tests [20]. The 95% confidence interval of this difference was estimated following the recommended method given by Altman et al. (19). The value 0.05 was used as threshold of statistical significance for all the statistical tests performed. All the analyses were done in the statistical software R version 3.6.1.

## Results

On May 1^st^, May 29^th^ and June 26^th^, respectively 40 (86.95%), 45 (97.82%) and 41 (89.13%) sub-Saharan African countries had a number of confirmed cases that was below the predicted number of 1000 cases for May 1^st^, and 10000 for both May 29^th^ and June 26^th^ (table 1). the population proportions of confirmed Covid-19 cases were significantly lower (p-value <0.05) than the projected proportions of cases. (table 2).

**Table 1.** Percentage difference between the projected and actual number of COVID-19 cases in sub-Saharan African countries on May 1^st^, May 29^th^, and June 26^th^

| Country | May 1<sup>st</sup> |  |  | May 29<sup>th</sup> |  |  | June 26<sup>th</sup> |  |  |
| --- | --- | --- | --- | --- | --- | --- | --- | --- | --- |
|  | Projected | Actual | Percent difference | Projected | Actual | Percent difference | Projected | Actual | Percent difference |
| Angola | 1000 | 27 | -97.3 | 10000 | 73 | -99.27 | 10000 | 212 | -97.88 |
| Benin | 1000 | 84 | -91.6 | 10000 | 218 | -97.82 | 10000 | 10147 | +1.47* |
| Botswana | 1000 | 23 | -97.7 | 10000 | 35 | -99.65 | 10000 | 125 | -98.75 |
| Burkina Faso | 1000 | 645 | -35.5 | 10000 | 847 | -91.53 | 10000 | 934 | -90.66 |
| Burundi | 1000 | 15 | -98.5 | 10000 | 42 | -99.58 | 10000 | 144 | -98.56 |
| Cabo Verde | 1000 | 121 | -87.9 | 10000 | 390 | -96.1 | 10000 | 1003 | -89.97 |
| Cameroon | 1000 | 1832 | +83.2* | 10000 | 5436 | -45.64 | 10000 | 12592 | +25.92* |
| CAR | 1000 | 64 | -93.6 | 10000 | 755 | -92.45 | 10000 | 3244 | -67.56 |
| Chad | 1000 | 73 | -92.7 | 10000 | 726 | -92.74 | 10000 | 863 | -91.37 |
| Comoros | 1000 | 1 | -99.9 | 10000 | 43 | -99.57 | 10000 | 272 | -97.28 |
| Congo | 1000 | 220 | -78 | 10000 | 571 | -94.29 | 10000 | 1204 | -87.96 |
| Côte d'Ivoire | 1000 | 1275 | +27.5* | 10000 | 2641 | -73.59 | 10000 | 8334 | -16.66 |
| DRC | 1000 | 572 | -42.8 | 10000 | 2832 | -71.68 | 10000 | 6410 | -35.9 |
| Equatorial Guinea | 1000 | 315 | -68.5 | 10000 | 1043 | -89.57 | 10000 | 1043 | -89.57 |
| Eritrea | 1000 | 39 | -96.1 | 10000 | 39 | -99.61 | 10000 | 144 | -98.56 |
| Eswatini | 1000 | 100 | -90 | 10000 | 279 | -97.21 | 10000 | 706 | -92.94 |
| Ethiopia | 1000 | 131 | -86.9 | 10000 | 831 | -91.69 | 10000 | 5175 | -48.25 |
| Gabon | 1000 | 276 | -72.4 | 10000 | 2431 | -75.69 | 10000 | 5087 | -49.13 |
| Gambia | 1000 | 12 | -98.8 | 10000 | 25 | -99.75 | 10000 | 43 | -99.57 |
| Ghana | 1000 | 2074 | +107.4* | 10000 | 7303 | -26.97 | 10000 | 15473 | +54.73* |
| Guinea | 1000 | 1495 | +49.5* | 10000 | 3553 | -64.47 | 10000 | 5211 | -47.89 |
| Guinea-Bissau | 1000 | 201 | -79.9 | 10000 | 1195 | -88.05 | 10000 | 1556 | -84.44 |
| Kenya | 1000 | 396 | -60.4 | 10000 | 1618 | -83.82 | 10000 | 5384 | -46.16 |
| Lesotho | 1000 | 0 | -100 | 10000 | 2 | -99.98 | 10000 | 20 | -99.8 |
| Liberia | 1000 | 141 | -85.9 | 10000 | 269 | -97.31 | 10000 | 681 | -93.19 |
| Madagascar | 1000 | 132 | -86.8 | 10000 | 656 | -93.44 | 10000 | 1829 | -81.71 |
| Malawi | 1000 | 37 | -96.3 | 10000 | 203 | -97.97 | 10000 | 960 | -90.4 |
| Mali | 1000 | 490 | -51 | 10000 | 1194 | -88.06 | 10000 | 2039 | -79.61 |
| Mauritania | 1000 | 8 | -99.2 | 10000 | 346 | -96.54 | 10000 | 3519 | -64.81 |
| Mauritius | 1000 | 332 | -66.8 | 10000 | 334 | -96.66 | 10000 | 341 | -96.59 |
| Mozambique | 1000 | 76 | -92.4 | 10000 | 233 | -97.67 | 10000 | 788 | -92.12 |
| Namibia | 1000 | 16 | -98.4 | 10000 | 22 | -99.78 | 10000 | 102 | -98.98 |
| Niger | 1000 | 719 | -28.1 | 10000 | 955 | -90.45 | 10000 | 1059 | -89.41 |
| Nigeria | 1000 | 1932 | +93.2* | 10000 | 8915 | -10.85 | 10000 | 22614 | +126.14* |
| Rwanda | 1000 | 243 | -75.7 | 10000 | 349 | -96.51 | 10000 | 850 | -91.5 |
| ST & P | 1000 | 16 | -98.4 | 10000 | 295 | -97.05 | 10000 | 392 | -96.08 |
| Senegal | 1000 | 933 | -6.7 | 10000 | 3348 | -66.52 | 10000 | 6233 | -37.67 |
| Seychelles | 1000 | 11 | -98.9 | 10000 | 11 | -99.89 | 10000 | 16 | -99.84 |
| Sierra Leone | 1000 | 124 | -87.6 | 10000 | 812 | -91.88 | 10000 | 1381 | -86.19 |
| South Africa | 1000 | 5647 | +464.7* | 10000 | 27403 | +174.03* | 10000 | 118375 | +1083.75* |
| South Sudan | 1000 | 35 | -96.5 | 10000 | 994 | -90.06 | 10000 | 1942 | -80.58 |
| Togo | 1000 | 116 | -88.4 | 10000 | 422 | -95.78 | 10000 | 588 | -94.12 |
| Uganda | 1000 | 81 | -91.9 | 10000 | 410 | -95.9 | 10000 | 821 | -91.79 |
| URT | 1000 | 480 | -52 | 10000 | 509 | -94.91 | 10000 | 509 | -94.91 |
| Zambia | 1000 | 106 | -89.4 | 10000 | 1057 | -89.43 | 10000 | 1497 | -85.03 |
| Zimbabwe | 1000 | 32 | -96.8 | 10000 | 132 | -98.68 | 10000 | 551 | -94.49 |

\*Percent difference greater than 1
\*\*Central African Republic
\*\*\*Democratic Republic of Congo
\*\*\*\*Sao Tome and Principe
\*\*\*\*\*United Republic of Tanzania

**Table 2.** P-value of the N-1 chi-square test comparing the projected and actual proportions of COVID-19 infection in African countries on May 1^st^, May 29^th^, and June 26^th^, along with the 95% confidence interval of the difference between these proportions

| Countries | May 1<sup>st</sup> |  | May 29<sup>th</sup> |  | June 26<sup>th</sup> |  |
| --- | --- | --- | --- | --- | --- | --- |
|  | p-value | 95% CI | p-value | 95% CI | p-value | 95% CI |
| Angola | <0.0001 | -0.00298 to -0.00294 | < 0.0001 | -0.03026 to -0.03015 | < 0.0001 | -0.02984 to 0.03113** |
| Benin | <0.0001 | -0.00761 to -0.00750 | < 0.0001 | -0.08085 to -0.08053 | < 0.0001 | 0.00099 to 0.16641 |
| Botswana | <0.0001 | -0.04180 to -0.04128 | < 0.0001 | -0.42432 to -0.42305 | < 0.0001 | -0.42049 to 0.43112** |
| Burkina Faso | <0.0001 | -0.00174 to -0.00166 | < 0.0001 | -0.04388 to -0.04369 | < 0.0001 | -0.04347 to 0.05240** |
| Burundi | <0.0001 | -0.00834 to -0.00823 | < 0.0001 | -0.08390 to -0.08359 | < 0.0001 | -0.08305 to 0.08547** |
| Cabo Verde | <0.0001 | -0.15917 to -0.15701 | < 0.0001 | -0.016931 to -0.017643 | < 0.0001 | -0.015817 to -0.016550 |
| Cameroon | <0.0001 | 0.00310 to 0.00317 | < 0.0001 | -0.01728 to -0.01710 | < 0.0001 | 0.00966 to 0.08521 |
| CAR*** | <0.0001 | -0.01951 to -0.01925 | < 0.0001 | -0.19179 to -0.19103 | < 0.0001 | -0.14030 to 0.27463** |
| Chad | <0.0001 | -0.00568 to -0.00560 | < 0.0001 | -0.05658 to -0.05634 | < 0.0001 | -0.05575 to 0.06625** |
| Comoros | <0.0001 | -0.11550 to -0.11416 | < 0.0001 | -0.01122 to -0.01167 | < 0.0001 | -0.01095 to -0.01141 |
| Congo | <0.0001 | -0.01426 to -0.01401 | < 0.0001 | -0.17121 to -0.17054 | < 0.0001 | -0.15975 to 0.20339** |
| Côte d'Ivoire | <0.0001 | 0.00101 to 0.00108 | < 0.0001 | -0.02798 to -0.02782 | < 0.0001 | -0.00641 to 0.06960** |
| DRC**** | <0.0001 | -0.00049 to -0.00047 | < 0.0001 | -0.00803 to -0.00798 | < 0.0001 | -0.00404 to 0.01835** |
| Equatorial Guinea | <0.0001 | -0.04932 to -0.04833 | < 0.0001 | -0.63928 to -0.63755 | < 0.0001 | -0.63928 to 0.78796** |
| Eritrea | <0.0001 | -0.02728 to -0.02693 | < 0.0001 | -0.28138 to -0.28044 | < 0.0001 | -0.27842 to 0.28654** |
| Eswatini | <0.0001 | -0.07812 to -0.07705 | < 0.0001 | -0.83870 to -0.83733 | < 0.0001 | -0.80197 to 0.92369** |
| Ethiopia | <0.0001 | -0.00076 to -0.00075 | < 0.0001 | -0.00799 to -0.00796 | < 0.0001 | -0.00422 to 0.01322** |
| Gabon | <0.0001 | -0.03283 to -0.03222 | < 0.0001 | -0.34080 to -0.33926 | < 0.0001 | -0.22156 to 0.67862** |
| Gambia | <0.0001 | -0.04113 to -0.04062 | < 0.0001 | -0.41332 to -0.41208 | < 0.0001 | -0.41258 to 0.41614** |
| Ghana | <0.0001 | 0.00342 to 0.00349 | < 0.0001 | -0.00876 to -0.00860 | < 0.0001 | 0.01751 to 0.08208 |
| Guinea | <0.0001 | 0.00369 to 0.00384 | < 0.0001 | -0.04926 to -0.04892 | < 0.0001 | -0.03664 to 0.11600** |
| Guinea-Bissau | <0.0001 | -0.04094 to -0.04026 | < 0.0001 | -0.44818 to -0.44663 | < 0.0001 | -0.42986 to 0.58799** |
| Kenya | <0.0001 | -0.00114 to -0.00111 | < 0.0001 | -0.01563 to -0.01555 | < 0.0001 | -0.00863 to 0.02866** |
| Lesotho | <0.0001 | -0.04697 to -0.04640 | < 0.0001 | -0.46743 to -0.46609 | < 0.0001 | -0.46659 to 0.46846** |
| Liberia | <0.0001 | -0.01711 to -0.01685 | < 0.0001 | -0.19274 to -0.19204 | < 0.0001 | -0.18460 to 0.21153** |
| Madagascar | <0.0001 | -0.00316 to -0.00311 | < 0.0001 | -0.03382 to -0.03367 | < 0.0001 | -0.02958 to 0.04279** |
| Malawi | <0.0001 | -0.00507 to -0.00500 | < 0.0001 | -0.05131 to -0.05111 | < 0.0001 | -0.04736 to 0.05740** |
| Mali | <0.0001 | -0.00256 to -0.00248 | < 0.0001 | -0.04358 to -0.04338 | < 0.0001 | -0.03942 to 0.05955** |
| Mauritania | <0.0001 | -0.02147 to -0.02120 | < 0.0001 | -0.20799 to -0.20723 | < 0.0001 | -0.13982 to 0.29118** |
| Mauritius | <0.0001 | -0.05306 to -0.05197 | < 0.0001 | -0.76067 to -0.75914 | < 0.0001 | -0.76012 to 0.81374** |
| Mozambique | <0.0001 | -0.00298 to -0.00294 | < 0.0001 | -0.03131 to -0.03119 | < 0.0001 | -0.02954 to 0.03458** |
| Namibia | <0.0001 | -0.03897 to -0.03848 | < 0.0001 | -0.39328 to -0.39208 | < 0.0001 | -0.39014 to 0.39817** |
| Niger | <0.0001 | -0.00119 to -0.00113 | < 0.0001 | -0.03745 to -0.03728 | < 0.0001 | -0.03702 to 0.04577** |
| Nigeria | <0.0001 | 0.00045 to 0.00046 | < 0.0001 | -0.00054 to -0.00051 | < 0.0001 | 0.00610 to 0.01584 |
| Rwanda | <0.0001 | -0.00590 to -0.00579 | < 0.0001 | -0.07466 to -0.07437 | < 0.0001 | -0.07080 to 0.08392** |
| ST and P***** | <0.0001 | -0.45143 to -0.44720 | < 0.0001 | -0.04343 to -0.04520 | < 0.0001 | -0.04298% to -0.04477 |
| Senegal | <0.0001 | -0.00045 to -0.00035 | < 0.0001 | -0.03986 to -0.03960 | < 0.0001 | -0.02264 to 0.09709** |
| Seychelles | <0.0001 | 0.00947 to 0.01074 | < 0.0001 | 0.01000 to 0.01038 | < 0.0001 | 0.09999 to 0.01037 |
| Sierra Leone | <0.0001 | -0.01106 to -0.01090 | < 0.0001 | -0.11542 to -0.11494 | < 0.0001 | -0.10830 to 0.14292** |
| South Africa | <0.0001 | 0.00781 to 0.00786 | < 0.0001 | 0.02928 to 0.02941 | < 0.0001 | 0.18262 to 0.21656 |
| South Sudan | <0.0001 | -0.00868 to -0.00856 | < 0.0001 | -0.08063 to -0.08028 | < 0.0001 | -0.07217 to 0.10687** |
| Togo | <0.0001 | -0.01076 to -0.01060 | < 0.0001 | -0.11592 to -0.11546 | < 0.0001 | -0.11391 to 0.12812** |
| Uganda | 0.17722* | -0.00202 to -0.00200 | < 0.0001 | -0.02101 to -0.02092 | < 0.0001 | -0.02011 to 0.02370** |
| URT***** | <0.0001 | -0.00088 to -0.00086 | < 0.0001 | -0.01592 to -0.01586 | < 0.0001 | -0.01592 to 0.01763** |
| Zambia | <0.0001 | -0.00490 to -0.00483 | < 0.0001 | -0.04875 to -0.04854 | < 0.0001 | -0.04636 to 0.06265** |

\*P-value greater than 0.05
\*\*95% Confidence interval containing 0
\*\*\*Central African Republic
\*\*\*\*Democratic Republic of Congo
\*\*\*\*\*Sao Tome and Principe
\*\*\*\*\*United Republic of Tanzania

On May 1^st^ and May 29^th^, all the confidence intervals of the difference between the projected and actual proportions of cases excluded 0. On June 26^th^, 41 (75.92%) of the confidence intervals of the difference between these proportions excluded 0.

Across all these dates, South-Africa always exceeded the predicted number and population proportion of Covid-19 infections.

## Discussion

This cross-sectional study comparing the projected and actual number of COVID-19 cases in the 46 sub-Saharan African countries found that on the dates specified, most countries did not meet the predictions in terms of number of COVID-19 cases. These results might be explained by the prompt response of African leaders, the distribution of COVID-19 high risk groups in sub-Saharan African countries, the disagreement between the statistical model parameters (or assumptions) and the reality, and the low testing and reporting capacity of sub-Saharan African countries.

From its first description [1], the COVID-19 infection is characterized by a rapid spreading ability. Human hosts of the virus are major carriers that can favor its transmission if they move from place to place. Therefore, the reduction of imported cases was one of the first important preventive measures against the spread of the COVID-19 infection. African leaders were quick to react; on March 11^th^ 2020 the WHO declared the COVID-19 outbreak a global pandemic [2], and as of March 15^th^, most African countries had closed their borders until further notice [21,22]. Thus, notwithstanding the harsh socioeconomic consequences, these results justify the positive impact of border closures towards limiting the spread of COVID-19. African public health authorities were also quick to implement and enforce hygiene sanitation measures. Finally, quarantine measures were imposed for several weeks in most countries [23,24].

The Centers for Disease Control and Prevention (CDC) identified persons at higher risk of COVID-19 infection as those of older age (above 65years) and/or those with severe medical conditions like chronic kidney disease (CKD), obesity, and immunocompromised states especially chronic stress/anxiety, type 2 diabetes mellitus (T2DM), and HIV/AIDS [25].

The higher risk associated with older age is related to the weakening of the immune system that occurs with aging [26,27]. Furthermore, children have been found to be at lower risk and to present milder symptoms of COVID-19 infection [28–30]. According to the United Nations Population Report [17], in 2019 there were 45 526 000 people aged 65 and older in Africa, which represents 3.5% of the total population. This is quite low compared to the United States (US) and Europe where they represent 16% and 18.8 % of the total population respectively [17]. Furthermore, the African population aged 0 to 14 years makes 40.55% of the inhabitants of the continent. Thus, the natural composition of the sub-Saharan African population might explain part of its resilience to the COVID-19.

The higher risk associated with CKD, obesity, and immunocompromised states that are chronic stress/anxiety, T2DM and HIV/AIDS is also linked to the weakening of the immune system that characterizes these pathologies [31–35]. Despite a relatively higher burden of HIV/AIDS [36] and CKD [37], the African population hosts lower proportions of individuals with chronic stress/anxiety [38], obesity [39] and T2DM [40] as compared to the US and Europe. Obesity is a risk factor for several pathologies [39] which in turn are also favoring conditions for the development of the COVID-19 infection; such a synergistic effect between obesity and these pathologies might account for part of the higher COVID-19 burden seen in the US and Europe as compared to Africa.

The difference observed between the projections and actual figures of the COVID-19 burden in Africa might also be explained by the fact that the epidemic parameters and /or assumptions used in the predictive model did not match with the reality on the ground. The epidemic parameters used considered that each case would produce an average of 2 additional cases [12], and that the average time between the onset of a case and the onset of a subsequent case infected by that case (serial interval) would be 4.7 days [13]. These parameters presented several limitations. First, they were estimated in the Chinese population which substantially differs from the sub-Saharan African population in many aspects that can fundamentally impact the spread of the COVID-19 infection. Furthermore, Abbot et al., did acknowledge the scarcity of data at the time they built their model and they further admitted that their results would be significantly impacted by the availability of new data [12]. In the same line, Bi et al., did point out that their study had “numerous limitations”, including the high risk of bias due to the multiplicity of data collection protocols, the fact that it was “impossible to identify every potential contact an individual has”, and the fact that asymptomatic travelers were missed [13]. Finally, for such predictions to come true, the living conditions should remain stable in order to agree with the statistical model parameters. However, the COVID-19 pandemic is an ever-changing affection which prevent predictive models from describing it accurately.

It is important to note that the actual figures might be low because all cases might not have been reported. Because of the novelty of the COVID-19 infection, testing kits had to be made from scratch. Countries with technical and infrastructural abilities were the first to dispose of COVID-19 tests. The African continent suffers from a substantial lack of high technical capacities that can produce such quality tests in sufficient amounts; therefore, the continent entirely relies on other countries to scale-up its testing abilities. Because of the high global burden of the COVID-19 infection, countries with testing capacities did put their populations’ interests first, leaving dependent countries like African ones on endless waiting lists. This, in addition to the overall weak health system (low financial capacity for purchasing tests; low man power capacity and communication infrastructures to ensure wide geographic coverage of testing teams or decentralization of testing laboratories) hampered the capacity of African countries to massively test their populations early, which might in part explains the low number of reported cases. For instance, the testing capacity measured by the number of tests performed per one million population is for instance 142325 in UK, compared to 672 in Nigeria [41]. Furthermore, African countries do not possess strong public health reporting systems, thus the number presented might not accurately describe current reality.

Finally, whether some specific individual or environmental factors have come into play by for instance reducing the virulence and spread of the virus, is still questioned. Research has suggested that vaccination with Bacillus Calmette-Guérin (BCG) can have protective effects against the COVID-19 infection [42], and BCG vaccination is compulsory at birth in almost all sub-Saharan African countries as part of national enlarged vaccination programs. High temperatures and humidity have also been suggested to be protective against the COVID-19 infection [43], and sub-Saharan African rests in high daily average temperatures and humidity percentages [44]. Research has also hinted at a protective effect of Vitamin-D against the COVID-19 infection [45]; sub-Saharan African countries are under high sun exposure year-round which confer higher levels of Vitamin-D to their populations, compared to Western countries [46]. Studies have also described that African populations have specific genetic characteristics and adaptations that have modified their susceptibility to various infections [47], thus leading scientists to hypothesize about a potentially stronger natural resistance of sub-Saharan African populations to the COVID-19 infection

Our study has three major limitations. First, there is a risk of ecologic fallacy since we used individual data to do our analyses but rather used population data to explain our findings. Second, statistical tests, p-values and confidence intervals carry their share of limitations and insufficiencies that need to be mentioned and accounted for. Finally, we used secondary data that were already collected, and therefore our study is exposed to the potential biases that might have arisen during the collection of such data.

## Conclusion

Our study is the first to provide tangible evidence of the fact that sub-Saharan Africa truly defied the dire predictions of the spread of the COVID-19 infections. Our study can inform African stakeholders about their continent’s unique advantageous characteristics that can form the ground for efficient and effective public health strategies against the COVID-19 pandemic, and beyond, against all the health hazards that deprive the continent from a much needed healthy population.

## Data Availability

The data can be accessible upon request to the corresponding author

https://apps.who.int/iris/handle/10665/332055

https://reliefweb.int/report/world/coronavirus-disease-covid-19-situation-report-130-29-may-2020

https://reliefweb.int/report/world/covid-19-disease-response-situation-report-21-20-26-june-2020

https://www.un.org/development/desa/publications/world-population-prospects-2019-highlights.html

## Contributors

DCFM and SPC conceived the project. DCFM drafted the manuscript, extracted the data, and performed the analyses. DCFM, AA, MYAN, MVK, SFM and SPC critically revised the manuscript for methodological and intellectual content. All authors approved the final version of this protocol.

